# How might postprandial ‘extras’ fatten? Simulation of a gastric mixing hypothesis

**DOI:** 10.1101/2022.05.17.22275194

**Authors:** David A. Booth

## Abstract

It was suggested that the ingestion of extra calories towards the end of or shortly after a meal might be especially fattening. That hypothesis was based on the intuition that mixing of the sugared drink and accompaniments while the stomach was emptying rapidly would delay the release of hunger less than the same intake in the hour before the next meal. This paper presents an examination of that mechanism by calculating the time course of gastric emptying with extra intake at different times after the meal. The output from these simulations confirmed that early further energy would delay the end of emptying less than later. However, within the parameters tested, the effect is not large. Fattening effects of calories after meals could arise by a variety of mechanisms that remain to be tested without disrupting daily life.

**“Highlights”:**

- Extra intake may be more fattening shortly after meals than later.
- That proposal was supported by theoretical calculations of gastric emptying.
- Hence the timing of energy intake well before a meal seems critical to its effect on weight.
- The eater’s own concepts of meal, snacks and drinks are key to research on weight control.

## Introduction

This brief paper presents the results of quantitative simulation of the emptying of the stomach after a meal. A small amount of extra energy is calculated to have been ingested at the end of the meal or after a delay. The question is how much this additional drink and/or food energy delays the slowing of gastric emptying to a rate at which the motivation to eat again is released (Booth & Mather, 1978).

Consuming food or a calorific drink within an hour before a meal is well known to reduce intake at that meal, regardless of types of food and their nutrient contents (e.g., Booth, 1981; Rolls, 2010). Clearly the extra energy ingested will increase the rate of gastric emptying at all later times and so may prevent hunger from arising by the usual time to eat the next meal. However, if the food is eaten soon after the previous meal, there will not be such a large relative increase in the already rapid rate of emptying. On these grounds, it was proposed that the earlier the extra energy enters the body, the less delay there is in the final phase of slow emptying and so the less likely it is that subsequent appetite is suppressed; hence there could be a greater increase in the day’s total intake of energy than from energy longer after the meal (Booth, 1988a). It seems that this is still the only quantitatively specified process proposed which triggers hormonal and metabolic mechanisms that make consumption of calories between meals particularly fattening (Chapelot, 2011).

Non-invasive studies have provided evidence that habitual consumption of a calorific drink or food between regular meals makes a substantial contribution to gain in weight (Bertius Forslund *et al*., 2005; Blair *et al*., 1989; Coakley *et al*., 1998; Kayman *et al*., 1990; Peneau *et al*., 2008). In the USA, this habit can be identified as “snacking” (Kayman *et al*., 1990). In the UK, however, “having a snack” can mean eating a light meal, for example at lunchtime (Chamontin *et al*., 2003; Drummond *et al*., 1996); the phrase “calories between meals” is widely understood there and less ambiguous than “a snack” (Booth *et al*., 2004). Indeed, “zero-calorie drink breaks” had been suggested as a first line of defence against unhealthy weight gain (Booth, 1988a).

There is no clear evidence that the source of the energy influences a fattening effect of the intake of energy early between meals -- starch, sugar, fat, alcohol or even protein. A fattening effect has been seen from sugar in sodas, often consumed between meals (e.g., Tordoff & Alleva, 1990). Fats in foods eaten at any time may be more fattening than carbohydrates because deposition of fatty acids from fat intake takes less energy than their synthesis from other ingested energy nutrients. This may explain an association of calories between meals and the eating of high fat foods as the most fattening practices (Booth *et al*., 2004) in the late 1980s English Midlands study (Blair *et al*., 1989). Certainly the eating of so-called “snack foods” regardless of timing has no association with weight gain (Conner & Norman, 1996). The hypothesis that eating soon after meals is especially fattening cannot be tested by dividing recorded bouts of intake at an arbitrary criterion of energy because such analyses disregard the timing of the small intake (McBride *et al*., 1990; Gatenby, 1997).

The proposal that extra energy intake shortly after a meal is especially fattening was informed by experience with a remarkably successful, physiologically realistic model of human meal patterns (Booth & Mather, 1978; Booth 1988b). However, simulations of the postulated effects of gastric mixing had not been run when the mechanism was mooted (Booth, 1988a). There has been no opportunity as yet to test the idea by measuring gastric emptying rates non-invasively after a meal, with additional energy intake at varying delays, let alone to extend such experiments to processes influenced by emptying. Hence this paper simply presents some estimates of the time course of rates of passage of food from the stomach, until the low rate is reached that was simulated to release hunger in the complete model (Booth & Mather, 1978; Booth, 1988b).

A sufficiently slow rate of passage of energy into the small intestine was hypothesised to be the ultimate physiological origin of the rise of the disposition to eat again because, with the exception of gastric distension of course, the strengths of the wide range of satiety signals depend on delivery of food to digestion and absorption. On that theory, relative failure of the extra food to delay the end of emptying is liable to advance the start of a search for food and to increase the amount eaten at the resultant meal. This lack of compensation for the extra energy consumed could be fattening if the intake of energy after that second meal was not reduced.

## Method

The time course of decrease in contents of the stomach was calculated in discrete steps of one minute, as in the original simulations (Booth & Mather, 1978). The formula used at each step was the amount in the stomach in kilocalories less 1.915 times the square root of that amount. That rate constant was one of the parameters in the published computer program that had been optimised to a person maintaining a constant weight of 75 kg (165 pounds) with a relatively sedentary pattern of physical activity (Booth & Mather, 1978). Gastric contents are not differentiated among energy nutrients because of evidence that, calorie for calorie, glucose, amino acids and fatty acids in the small intestine are equally effective at slowing gastric emptying (Hunt & Stubbs, 1975).

Gastric emptying was simulated as decelerating in accord with the square root of its present contents, for the following reasons. The emptying of meals on mixtures of solids and liquids clearly slows as the contents of the stomach become smaller. However, a genuinely exponential function may be limited to purely liquid loads. The emptying of a solid can be linear until the very last stages because of the extra processes of mechanical and digestive reduction of lumps of food to a liquid suspension before they can pass the pylorus. Data on gastric contents in rats after a meal on maintenance diet fitted a square root equation better than an exponential or a straight line (Newman & Booth, 1981). The curvature of a square root function is less than that of an exponential, hence averaging with linear emptying. The square root also is a theoretical function for emptying a cylinder, a shape to which the human stomach approximates (Hopkins, 1966). The square-root equation (Booth, 1988b; Booth & Mather, 1978) was used subsequently to model both the regulation of blood glucose by secretion of insulin in response to meals (Campfield, Smith & Fung, 1982) and also the effects of social environment and familial genetics on the amounts eaten in meals recorded in diaries (de Castro, 1999; de Castro *et al*., 1986).

Calculations were carried out in Microsoft Excel and graphs constructed in SigmaPlot.

## Results

In a 75-kg person who is not gaining weight at a moderate level of physical activity (Booth & Mather, 1978), an initial contents of 1000 kcal at the end of a meal was simulated to pass from the stomach to the small intestine at a decreasing rate that reached one kilocalorie per minute (a gastric content of 27 kcal) at about 4.5 hours (Figure 1, lowest curve). In the original simulation, gastric emptying at 1 kcal/min was taken to be the threshold for onset of motivation to have another meal. Hence, with a breakfast ending with that amount in the stomach at 7:30 a.m., this result corresponds to the person simulated becoming hungry for a lunch at 12 noon.

**Figure 1.**
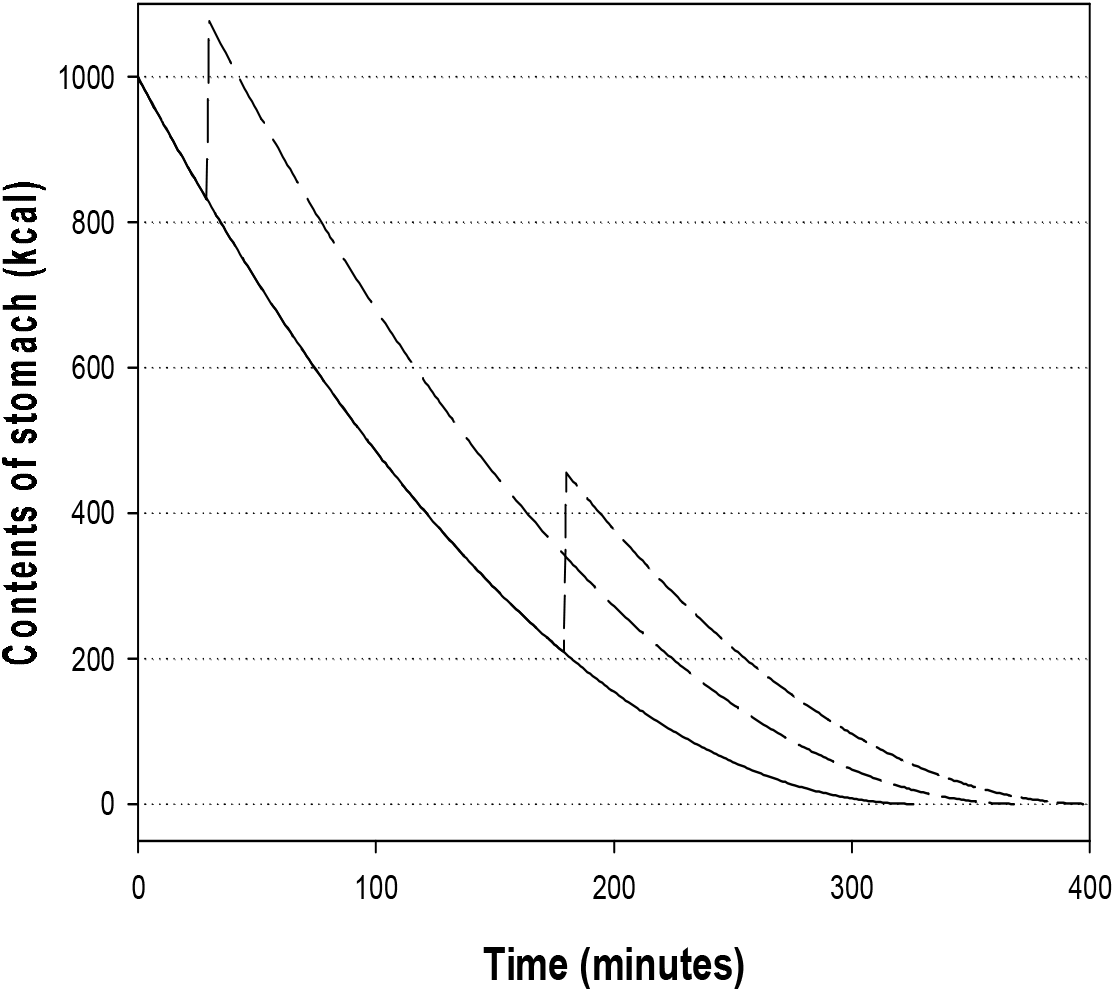
Simulated emptying from the stomach of a 1000-kcal residue from a meal (lowest curve) with intake of a further 250 kcal either 30 minutes after the meal (middle curve) or at 3 hours (highest curve after 180 minutes).

When a further 250 kcal was consumed within half an hour of the end of breakfast, the decrease of emptying rate to 1 kcal/min was delayed by about 40 minutes (Figure 1, middle curve). However, when the same amount of energy was ingested 3 hours after the meal (at 10:30 a.m. on the above schedule), the decrease in emptying rate to 1 kcal/min was delayed by over 70 minutes, taking nearly 6 hours, to 1:20 p.m. from a 7:30 a.m. end of breakfast (Figure 1, top curve). That is, according to this simulation, the energy content of the extra intake delays hunger considerably longer when it is eaten at a time closer to the next meal than to the previous meal. Hence the extra energy intake longer after a meal is likely to be compensated by eating less at the next meal – for example, at a lunch started between 12 noon and 1:15 p.m. (compare Kissileff *et al*., 2008)

Those simulations were repeated for an extra intake of 50, 100 or 250 kcal, taken at one of the half-hour intervals from at the end of the meal to 30 minutes before readiness for the next meal without the extra intake (Figure 1). With the 50- and 100-kcal extra (including sugar drinks), intakes at 0 or 30 min differed negligibly in their delaying of hunger (Figure 2). An extra of 100 kcal up to 2 hours after the meal would still let the simulated person become hungry less the 40 min later than without the extra, i.e., by 12:40 after a 7:30 a.m. breakfast. However, when that size of extra intake is taken 4 hours after the meal (e.g., at 11:30 a.m. after a 7:30 a.m. breakfast), the delay of hunger increases by over half an hour (well past 1 p.m.).

**Figure 2.**
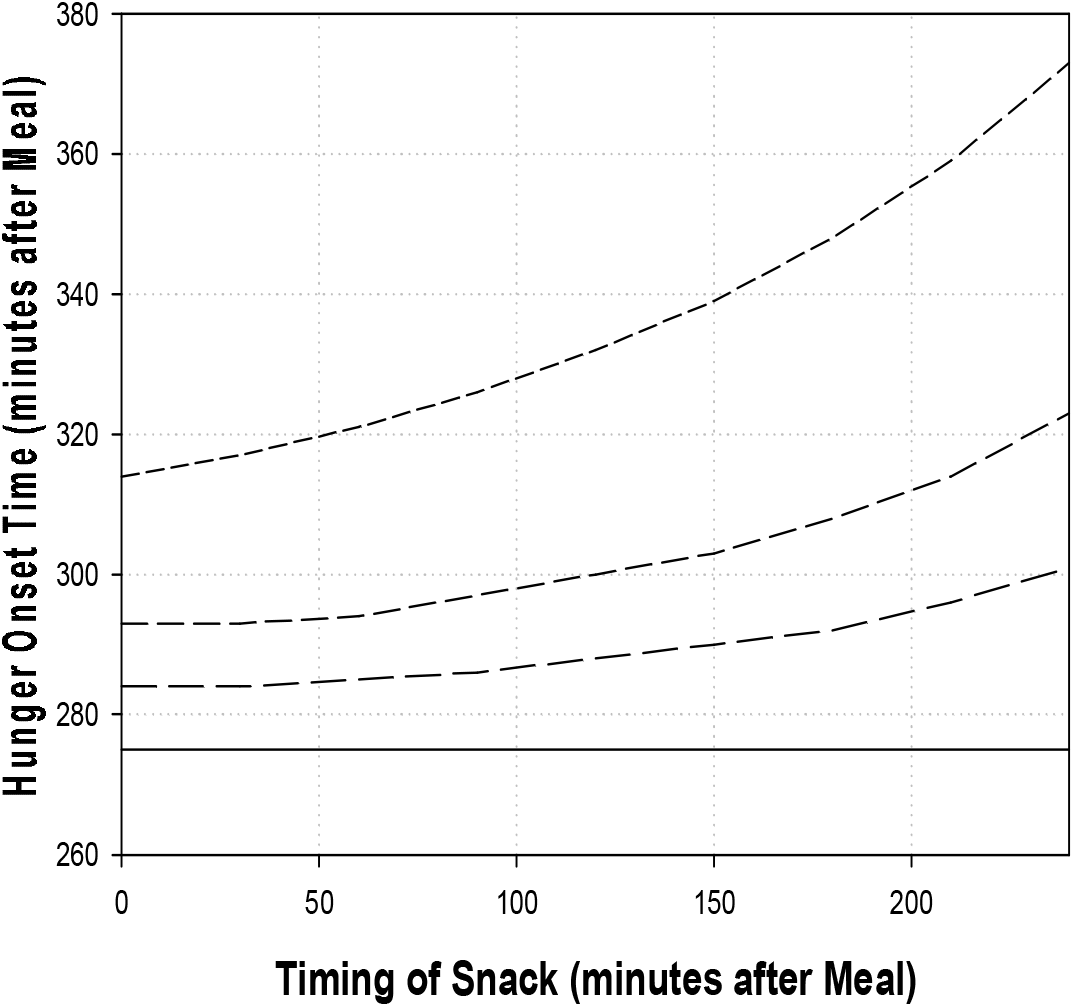
Increasing postponement of onset of hunger with longer delay of extra intake (50, 100 or 250 kcal) after the meal simulated in Figure 1 leaves 1000 kcal in the stomach. Tested points at 30-min intervals are connected by straight lines.

To sum up, the simulated decrease in contents of the stomach after a meal reached a very slow rate sooner when there was extra intake at the end of the meal than if the same intake occurred a few hours later. Hence, whatever the timing of the next meal, the earlier that any amount of extra energy is ingested, the less will be the physiological suppression of appetite and the more is liable to be eaten at the next meal.

## Discussion

### Earlier extras could be more fattening

Deceleration of gastric emptying was the basis of the intuition that habitual extra intake of energy more than an hour or so before a meal could be especially fattening. The thought was that mixing of the extra calories from a food or a drink with the previous meal during the more rapid phase of gastric emptying would make relatively little difference to the timing of the rise of hunger and hence the amount eaten at a socially fixed time. Another qualitative intuition with some of the same implications is that, the later the extra, the less of it will have emptied from the stomach whenever there is next some access to food.

Calculations on a plausible formula for the slowing of gastric emptying in the interval between meals confirmed that an extra later after a meal should generate more physiological signals of satiety than an earlier extra would. While emptying proceeds at a substantial rate, substrates for digestion in the small intestine continue to be replenished; this maintains chemical stimulation of innervated receptors in the wall of the digestive tract, the secretion of centrally active gut hormones and the absorption of substrates that sustain the supply of glucose and correlative signals to the brain.

These simulations also supported the original recognition that extra intake immediately following a meal could be especially fattening, e.g. chocolate, cream and/or alcohol in or with a postprandial drink (Booth, 1988a).

Furthermore, the hypothesised mechanism might implicate energy-rich desserts, and large or second portions of pudding. This extrapolation must be tentative because gastric emptying can be much faster during meals, e.g. because of ‘dumping’ into an empty duodenum (Hunt & Stubbs, 1975; cp. Kaplan, Spector & Grill, 1992, and Moran, Knipp & Schwartz, 1999), as well as a different pattern of secretion of the hormones implicated in states of satiety (Steiner, Meyer-Gerspach & Beglinger, 2012). For these reasons, research into the fattening effects of larger portions should always distinguish among timings later and earlier within meals.

### Further tests for fattening effects of calories between meals

It is increasingly recognised that unhealthy snacking is not the eating of small amounts or of packet foods or ‘empty calories’ but repeated intake of any calorific food or drink away from meals (Chapelot, 2011). Full attention has yet to be paid to two further specifics about any fattening effect of an increase in such habitual consumption of calories between meals.

First, closer attention needs to be paid to the thermodynamics by which the snacking could increase daily energy intake. Many have pointed out that, to the contrary, an increase in the frequency of smaller intakes might result in less total intake over a given period, thus reducing weight by a step. The energy-exchange mechanism for such an effect could be stronger compensatory influences from occasion to occasion of intake. The increase in rate of energy intake (average amount per period) considered here could come from worse compensation, because of too much delay between the snack and the next meal to maintain processes of satiation driven by continued delivery of energy nutrients to the duodenum.

The present results indicate that the time since the previous meal is crucial as well. Initially rapid postprandial gastric emptying implies that extra energy sooner after the previous meal is less likely to be compensated, whenever the next meal is eaten. Hence the recording of meal-to-snack times is as least as important as the recording of snack-to-meal times. Eating in the 90 minutes after a meal certainly should not be excluded from the definition of a snack if hypothesised mechanisms of fattening are to be tested (Viskaal -- van Dongen *et al*., 2010).

Secondly, whether or not invasive research designs are used, investigators need to distinguish between what serves as a meal from the calories between meals, i.e. the extra intake or the snack. A starting point is to measure intake that is not in meals identified as such by the eater in the local culture’s own terms (Blair *et al*., 1989; Booth & Booth, 2011; Laguna-Camacho *et al*., 2008). Such data can then yield the timings of intakes relative to both preceding and following meals, as well as those between-meal and end-of-meal intakes’ contents of energy nutrients and other potentially satiating agents.

## Data Availability

All data produced in the present work are contained in the manuscript.

